# Spatial and epidemiological drivers of *P. falciparum* malaria among adults in the Democratic Republic of the Congo

**DOI:** 10.1101/2020.01.28.20018978

**Authors:** Molly Deutsch-Feldman, Nicholas F. Brazeau, Jonathan B. Parr, Kyaw L. Thwai, Jérémie Muwonga, Melchior Kashamuka, Antoinette K. Tshefu, Ozkan Aydemir, Jeffrey A. Bailey, Jessie K. Edwards, Robert Verity, Michael Emch, Emily W. Gower, Jonathan J. Juliano, Steven R. Meshnick

## Abstract

**Background:** Malaria remains a significant public health problem in sub-Saharan Africa. Adults are frequently infected and may serve as a reservoir for further transmission, yet we know relatively little about risk factors for adult infections. In this study, we assessed malaria risk factors amongst adults using samples from the nationally representative, cross-sectional 2013-2014 Demographic and Health Survey (DHS) conducted in the Democratic Republic of Congo (DRC). We further explored differences in risk factors by urbanicity.

**Methods:** *Plasmodium falciparum* infection was determined by polymerase chain reaction (PCR). Covariates were drawn from the DHS to model individual, community, and environmental level risk factors for infection. Additionally, we used deep sequencing data to estimate the community-level proportions of drug resistant infections and included these estimates as potential risk factors. All identified factors were assessed for differences in associations by urbanicity.

**Results:** A total of 16,126 adults were included. Overall prevalence of malaria was 30.3% (SE = 1.1) by PCR; province-level prevalence ranged from 6.7-58.3%. Only 17% of individuals lived in households with at least one bednet for every two people, as recommended by the World Health Organization. Protective factors included increasing within-household bednet coverage (PR = 0.85, 95% CI = 0.76 - 0.95) and modern housing (PR = 0.58, 95% CI = 0.49 - 0.69). Community level protective factors included: increased average education and wealth (PR = 0.77, 95% CI = 0.65-0.91; PR - 0.84, 95% CI = 0.80 - 0.89). Education, wealth, and modern housing showed protective associations in cities but not in rural areas.

**Conclusions:** The DRC continues to suffer from a high burden of malaria; interventions that target high-risk groups and sustained investment in malaria control are sorely needed. Differences in risk factors by urbanicity may be due to differences in transmission intensity or access to resources.

## Introduction

Malaria remains an important cause of morbidity and mortality in the Democratic Republic of the Congo (DRC), which is home to 11% of all cases globally^1^. Understanding malaria transmission in the DRC is critical for furthering efforts to eliminate malaria in sub-Saharan Africa. In addition to the high disease burden, there is evidence that the DRC acts as a bridge of transmission, connecting parasites from East and West Africa^2–4^. To combat transmission and reduce disease burden, it is important to determine risk factors for infection. These factors can be used to identify individuals or groups more likely to become infected and, therefore, more likely to benefit from programmatic interventions. Adult infections remain understudied, as most malaria deaths occur amongst children^5,6^. However, adults are frequently infected, often asymptomatically, and may serve as a reservoir for transmission^7,8^. Yet, we know very little about risk factors for infection in adults. In this study, we assessed malaria risk factors amongst adults using data from over 16,000 participants from the nationally representative 2013-2014 Demographic and Health Survey (DHS) conducted in the DRC, the largest and most recent health survey conducted in the country.

A small number of studies performed in the DRC have identified important risk factors for infection^4,9,10^. These studies showed that increasing age, wealth, and individual bed-net use are protective, as are increasing community level bed-net use, lower average temperature and higher altitude^9–12^. However, these studies included only children under the age of five^9,10^, or were small and geographically limited^11,12^. A study of adults in the DRC conducted using data from the 2007 DHS identified several risk factors, such as younger age, male sex, and lower individual and community level wealth^13^; however, no nationally representative risk factor studies of adults have been conducted since.

In this study we evaluated current individual, household, community, and environmental risk factors for malaria amongst adults. Additionally, because increasing antimalarial drug resistance is a growing concern in the DRC, we sought to understand the relationship between community-level resistance and infection prevalence^14,15^. Similarly, the association between increasing antimalarial use and malaria prevalence amongst adults has not been studied in the DRC. Understanding the role of these factors is critical for determining drivers of malaria infection. The different scales of the factors demonstrate the various levels at which malaria interventions can be targeted.

This study builds on previous work in the DRC by evaluating the role of urbanization on risk factors for infection. Understanding the effect of urbanization on malaria transmission is a critical part of intervention planning^16–18^. Unlike other infectious diseases, which thrive in cities due to increased population density, malaria transmission is often lower in urban areas as compared to rural areas^19,20^. This is due to several factors, including reduced vector populations, lower biting rates, and better access to therapeutics^19^. As a result, the effects of malaria control programs have been shown to differ between cities and rural areas. Multiple studies conducted in Nigeria and Benin found that bednet ownership and use were higher amongst rural populations than urban^21–23^. Past work conducted in Liberia determined that the effects of bednets differed between the settings, with increasing community level bednet coverage being protective in urban areas but displaying no effect in rural areas^24^. We explored this relationship in the DRC by assessing whether the effects of various risk factors for infection differed between individuals in urban versus rural areas.

The findings from this study can be used to identify individuals and communities at higher risk for malaria infection in the DRC. They also shed light on differences in the epidemiology of malaria between urban and rural settings.

## Methods

### Study population

The data from this study are drawn from the 2013-2014 Demographic Health Surveys (DHS) conducted in the DRC. The DHS Program, run by USAID in conjunction with local governments, carries out periodic cross-sectional surveys in over 90 countries^25^. In the DRC, the DHS was conducted by the coordinated efforts of several governmental ministries. Using a nationally-representative randomized cluster sampling method, sampling cluster sites were first selected from a map of enumerated areas across the country with the probability of selection proportional to the size (number of households) of the cluster^26^. Next, households were randomly selected for inclusion from within clusters^27^. Cluster sizes differed between urban and rural areas with approximately 5 – 15 more households included in rural areas^27^. To account for this nested cluster sampling strategy when analyzing the data, each cluster was assigned a sampling weight. DHS survey conductors visited selected households, obtained consent, and administered an extensive questionnaire covering a broad range of topics including nutrition, education, health history, and infectious diseases status^26^. Blood samples were collected on filter paper for HIV testing; a second blood spot was also shipped to The University of North Carolina for molecular testing of malaria.

A total of 18,257 adults from 12,549 households were included and asked to provide blood samples. This included 9,601 women (ages 15-49) and 8,656 men (ages 15-59). In addition, 9,790 children (ages 6 months to 5 years) were part of the survey. Although they were not included in this analysis, they have been examined previously^9,14,28^.

### Risk factor selection

Potential risk factors *P. falciparum* infection were determined by consulting previous studies and based on biological plausibility. We selected individual, household, and cluster (community) level risk factors that have demonstrated associations with malaria risk ^9,10,13^. Individual factors included age, biological sex, HIV infection status, education, and wealth index. The DHS Program calculates wealth index as a composite variable based on each household’s assets and housing materials^29^. Using Principal Components Analysis (PCA), participants were then grouped into wealth quintiles: poorest, poor, middle, rich, and richest^29^. We also assessed bednet use, which was determined by whether the individual reported sleeping under a long lasting insecticide net (LLIN) the previous night. At the household level, we constructed a “net ratio” variable by dividing the number of total nets per household by the number of individuals within the household. This was then dichotomized into ratios of less than 0.5 versus those of 0.5 or higher (ie: at least one net for every two household members). We also created a composite “housing materials” variable, dichotomized as either traditional or modern, based on data from the roof, wall, and floor material variables (further details are available in the Supplemental Text)^30^. Cluster level factors included proportion reporting LLIN use, median wealth index, and median education level. We also assessed average annual precipitation and temperature. Data were drawn from the DHS questionnaire as well as from the DHS Program, which collects a range of environmental and geological data^31^.

### PCR diagnosis

The primary outcome for this study was *P. falciparum* malaria infection as determined by real-time PCR. Whole blood collected by finger-prick was spotted onto Whatman filter paper, dried at ambient temperature, and initially stored with dessicant at -80C in Kinshasa until punching (one 6mm punch per subject) and shipment to UNC. DNA was extracted from the blood spots using a Chelex extraction assay and used for PCR testing^32^. The PCR assay detects the *P*.*falciparum* lactate dehydrogenase gene (*pfldh*) with a limit of detection of 5-10 parasites/mL^33,34^. The human beta-tubulin (HumTuBB) gene was used as a positive control, and any samples that failed to amplify HumTuBB were excluded. The duplexed *pfldh* and HumTuBB quantitative PCR assay was performed using reaction conditions, primers, and quality control measures for high-throughput PCR exactly as previously described^35^. All samples were run in duplicate. Samples that amplified *pfldh* in only a single replicate were considered negative if the cycle threshold value was higher than 38. All laboratory assays were completed at the University of North Carolina at Chapel Hill.

### Genetic analyses

Community-level drug resistance was determined using second-generation sequencing data obtained using molecular inversion probes (MIPs) that target known molecular markers of resistance to antimalarial drugs^14^. MIPs are a technology for obtaining highly multiplexed deep sequencing data that have recently been applied to *Plasmodia* species^14^. Using previously generated sequencing data from 1,065 children enrolled in the 2013-14 DRC DHS^14,15^, we assessed single nucleotide polymorphisms (SNPs) of the *pfdhps* (A437G, K540E, and A581G) and *pfcrt* (K76T) genes known to be associated with sulfadoxine-pyramethamine (SP) and chloroquine resistance, respectively^14,36–38^. We used these individual-level data to estimate cluster-level prevalence for each SNP using the *PrevMap* package in R, which fits a spatial model using a Gaussian Process^39^. We fit the model to generate estimates of the underlying allele frequency distribution using maximum likelihood and running 10,000 simulations. Further model details are available in the Supplemental Text.

### Urban/rural classification

Potential misclassification of urbanicity is a concern in large surveys with complex sampling frames^40^. In order to minimize such misclassification, we conducted a PCA incorporating variables with a demonstrated relationship to urbanicity^31,41,42^. These variables were: degree of built environment, nighttime lights, total population as of 2014, population density as of 2014, and estimated travel time to the nearest city. The DHS collects these data from a variety of remote sensing and modeling databases including: WorldPop, the Joint Research Centre, and the Climate Research Unit at the University of East Anglia^31,43–45^. The DHS collects these variables for each cluster; all variables were scaled and log transformed for the analysis. From the PCA, we extracted the values for the first principal component (PC) for each cluster, and generated a new urban/rural variable by dichotomizing the PC1 value at the 75% percentile (e.g., urbanicity is represented by the upper quartile of PC1). The new measure of urbanicity was used to assess differences in risk factors between urban and rural areas.

### Modeling and statistical analyses

All analyses were performed using the R statistical platform v. 3.5.2^46^. We assessed the relationship between each risk factor and PCR detectable malaria infection using bivariate log-binomial regression models (i.e. not adjusted for other variables) fit using generalized estimating equations to estimate prevalence ratios. We used bivariate models as the aim was to assess marginal associations, not causal relationships. To account for the cluster sampling method, we included the DHS sampling weights using the *survey* package in R^47^. Each identified risk factor was then further assessed to determine if the association with malaria prevalence differed by urban/rural status by including the urban variable and an interaction term between each factor and the urban variable in the model. In general, variables were included in the model in the form that the data were collected (i.e.: categorical variables kept as categorical), though some continuous variables were scaled and proportions were logit transformed. As done previously for MIP genetic data, we generated spatially smoothed prevalence and standard error estimates using the *PrevMap* package (**Supplemental Text**)^39^.

In addition to the risk factor modeling we conducted several subsequent analyses. As the DHS asks multiple questions regarding bednet use, we conducted a sensitivity analysis to compare various coding methods for net use. Further details are presented in the Supplementary Text. We also assessed whether the association between individual net use and malaria risk differed across categories of overall cluster malaria prevalence. For this analysis we used previously published data from children included in the 2013-2014 DHS to determine community level prevalence in order to avoid including the outcome data of adult infections from 2013 in determining cluster level prevalences^9,28^.

### Patient and Public Involvement

This research was conducted without patient involvement

## Results

Our analysis included 16,126 adults from the original 18,257 initially selected for inclusion in the DHS. Individuals were sampled from 533 geographically dispersed clusters; GPS data were missing for 44 clusters, resulting in 489 clusters for analysis (**Figure 1**). After final data processing, 16,363 individuals had complete PCR and DHS covariate data. Two-hundred and thirty-seven individuals were not considered “du jure” (members of the sampled household, rather than visitors who slept in the household the previous night^48^) and, thus, were not included in the analysis, resulting in a final data set of 16,126 individuals (**Figure 2**). Overall PCR prevalence of *P. falciparum* infection was 30.3% (SE = 1.1). The results of the *PrevMap* analysis demonstrated the high spatial heterogeneity of infection; community prevalence estimates ranged from 0-76% (**Figure 3**). The map of the model standard errors indicated low variance in the estimates across the DRC (**Figure S1**). Prevalence estimates by province ranged from 6.7% in Nord-Kivu to 58.3% in Bas-Uele (**Supplementary Table 1**).

**Table 1:**
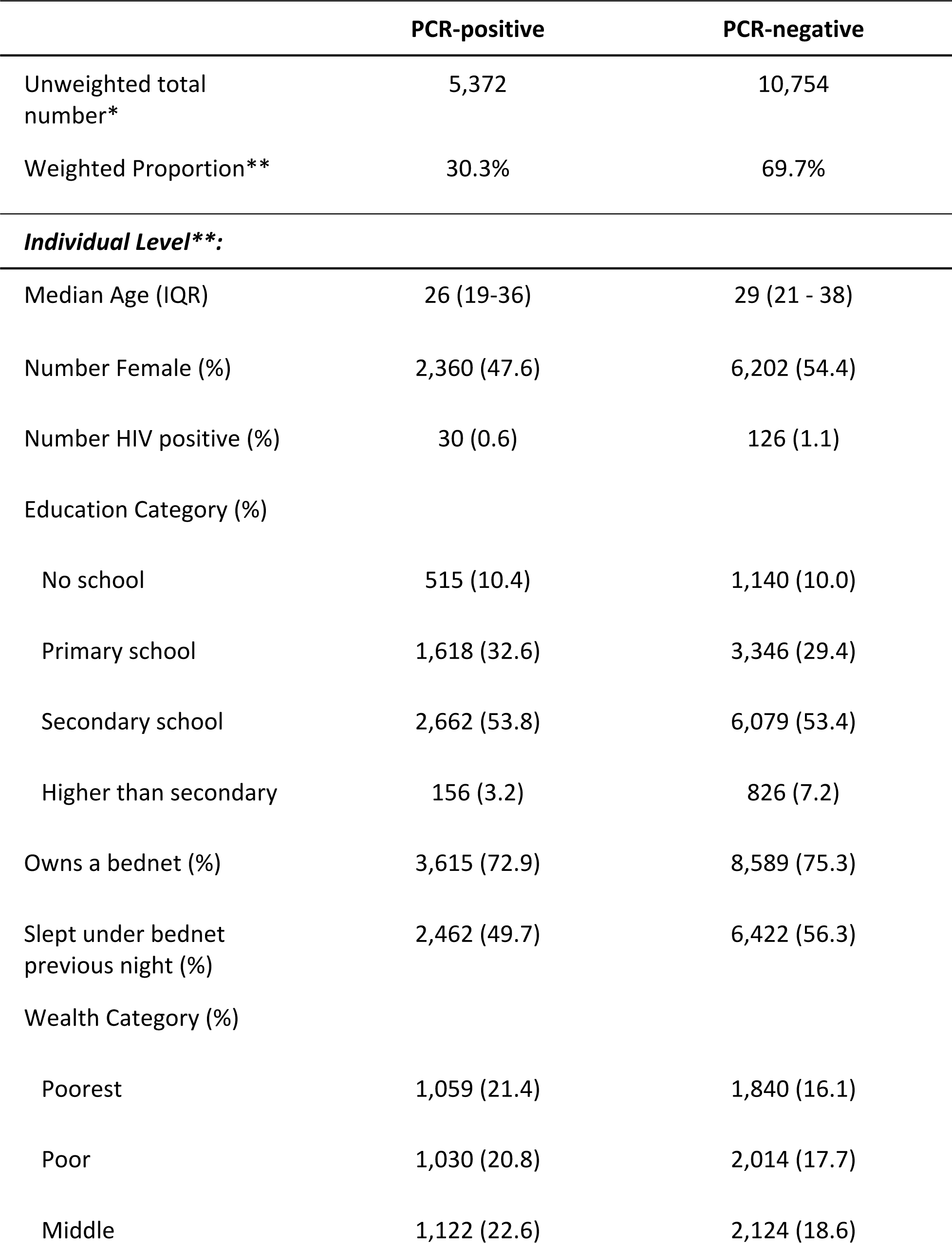
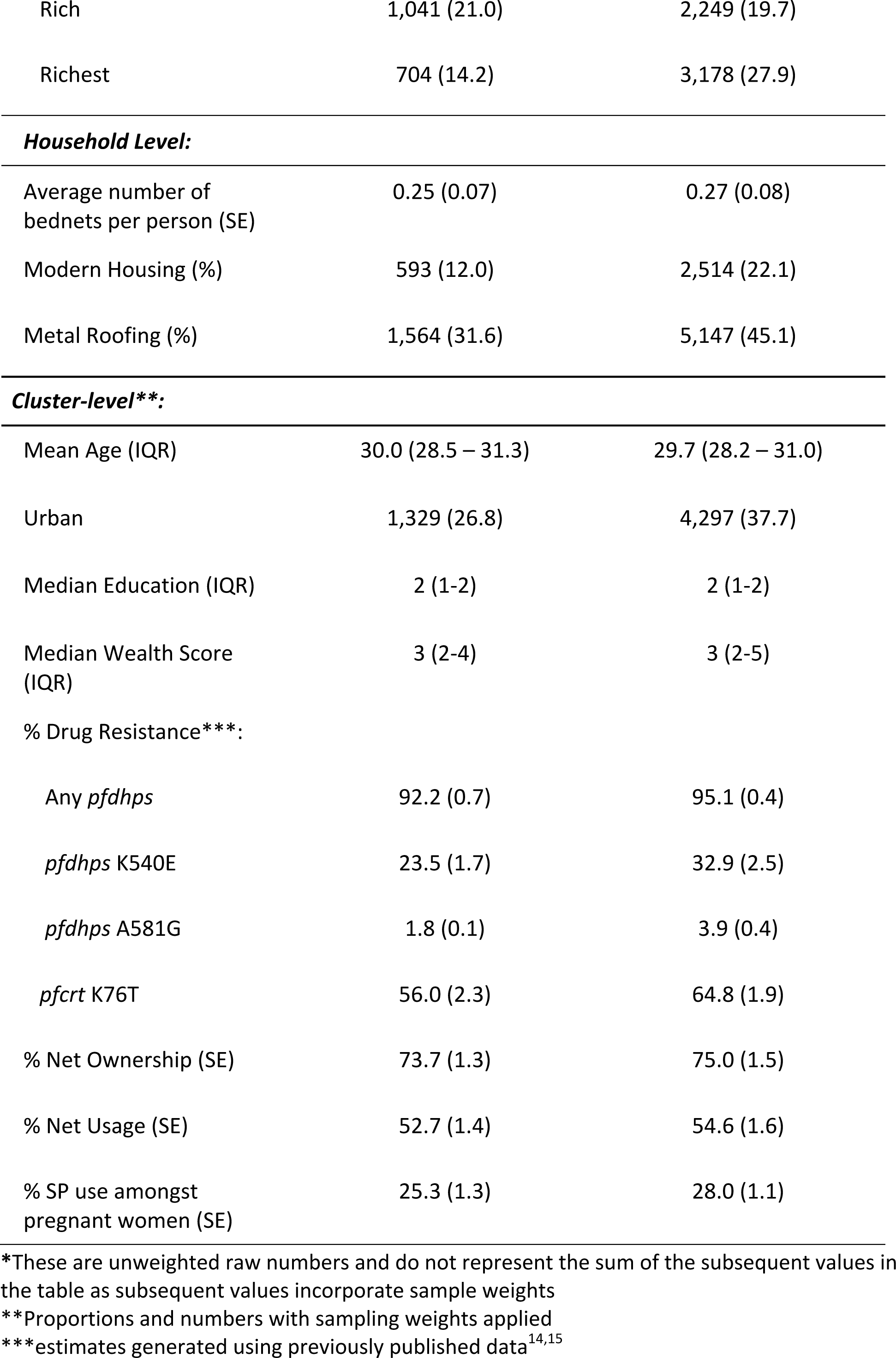
Descriptive statistics of the study population by *P. falciparum* PCR status

|  | PCR-positive | PCR-negative |
| --- | --- | --- |
| Unweighted total number* | 5,372 | 10,754 |
| Weighted Proportion** | 30.3% | 69.7% |
| ***Individual Level\*\*:*** |  |  |
| Median Age (IQR) | 26 (19-36) | 29 (21 - 38) |
| Number Female (%) | 2,360 (47.6) | 6,202 (54.4) |
| Number HIV positive (%) | 30 (0.6) | 126 (1.1) |
| Education Category (%) |  |  |
| No school | 515 (10.4) | 1,140 (10.0) |
| Primary school | 1,618 (32.6) | 3,346 (29.4) |
| Secondary school | 2,662 (53.8) | 6,079 (53.4) |
| Higher than secondary | 156 (3.2) | 826 (7.2) |
| Owns a bednet (%) | 3,615 (72.9) | 8,589 (75.3) |
| Slept under bednet previous night (%) | 2,462 (49.7) | 6,422 (56.3) |
| Wealth Category (%) |  |  |
| Poorest | 1,059 (21.4) | 1,840 (16.1) |
| Poor | 1,030 (20.8) | 2,014 (17.7) |
| Middle | 1,122 (22.6) | 2,124 (18.6) |
| Rich | 1,041 (21.0) | 2,249 (19.7) |
| Richest | 704 (14.2) | 3,178 (27.9) |
| ***Household Level:*** |  |  |
| Average number of bednets per person (SE) | 0.25 (0.07) | 0.27 (0.08) |
| Modern Housing (%) | 593 (12.0) | 2,514 (22.1) |
| Metal Roofing (%) | 1,564 (31.6) | 5,147 (45.1) |
| ***Cluster-level\*\*:*** |  |  |
| Mean Age (IQR) | 30.0 (28.5 – 31.3) | 29.7 (28.2 – 31.0) |
| Urban | 1,329 (26.8) | 4,297 (37.7) |
| Median Education (IQR) | 2 (1-2) | 2 (1-2) |
| Median Wealth Score (IQR) | 3 (2-4) | 3 (2-5) |
| % Drug Resistance***: |  |  |
| Any *pf dhps* | 92.2 (0.7) | 95.1 (0.4) |
| *pf dhps* K540E | 23.5 (1.7) | 32.9 (2.5) |
| *pf dhps* A581G | 1.8 (0.1) | 3.9 (0.4) |
| *pf crt* K76T | 56.0 (2.3) | 64.8 (1.9) |
| % Net Ownership (SE) | 73.7 (1.3) | 75.0 (1.5) |
| % Net Usage (SE) | 52.7 (1.4) | 54.6 (1.6) |
| % SP use amongst pregnant women (SE) | 25.3 (1.3) | 28.0 (1.1) |

\*These are unweighted raw numbers and do not represent the sum of the subsequent values in the table as subsequent values incorporate sample weights
\*\*Proportions and numbers with sampling weights applied
\*\*\*estimates generated using previously published data<sup>14,15</sup>

**Figure 1:**
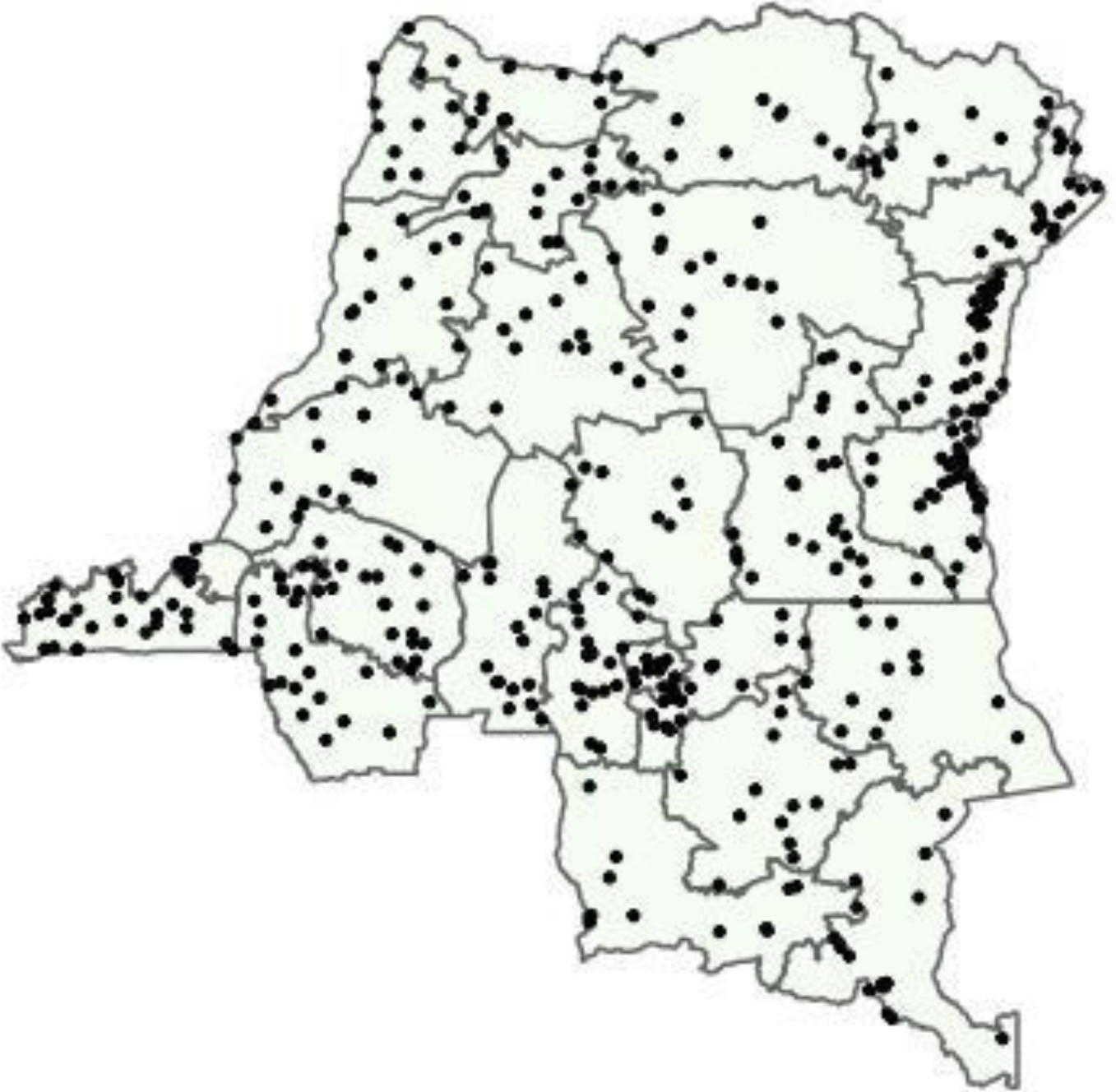
Sites of 2013-2014 DHS sampling clusters.

**Figure 2:**
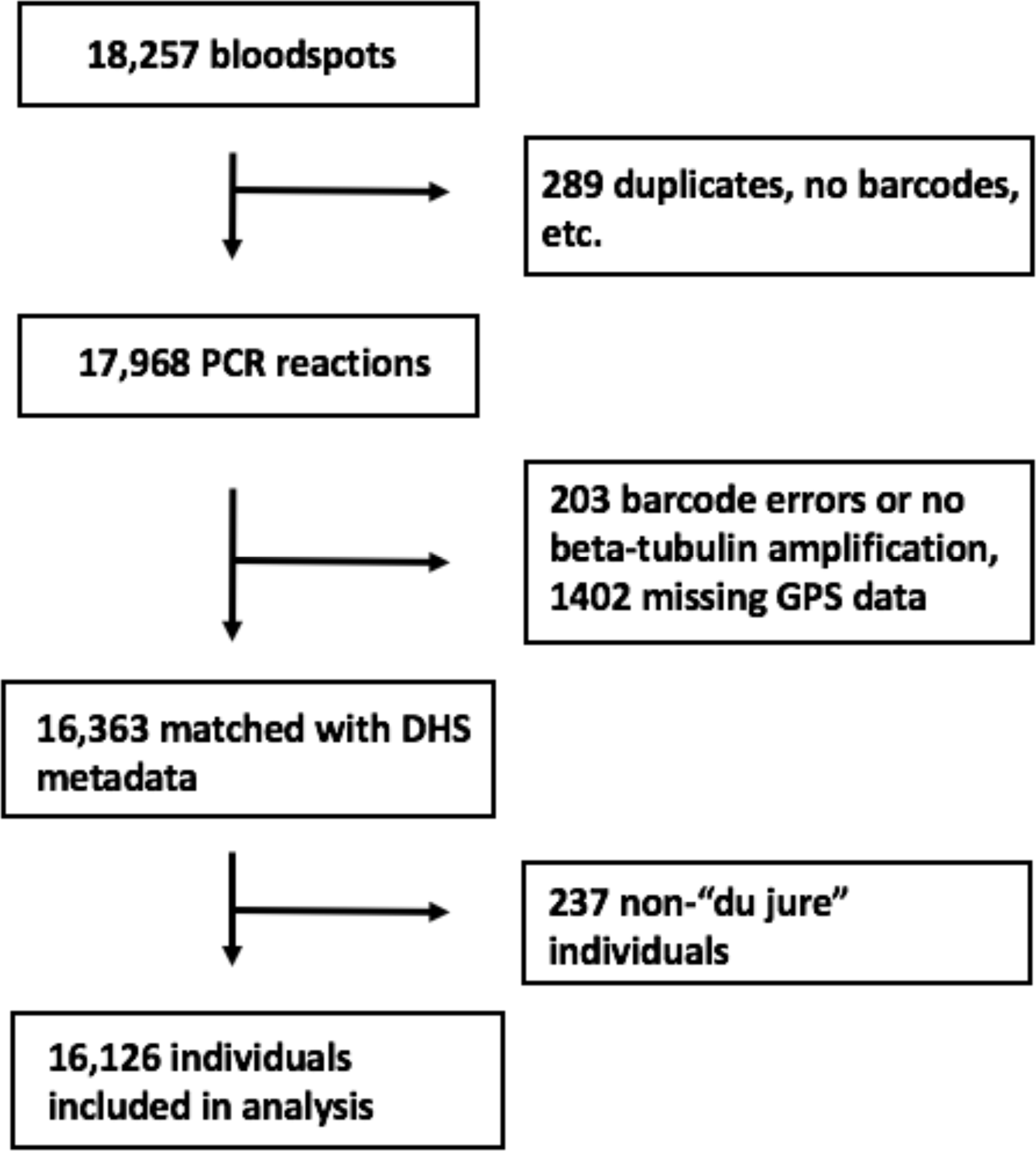
Flowchart of samples included in analysis.

**Figure 3:**
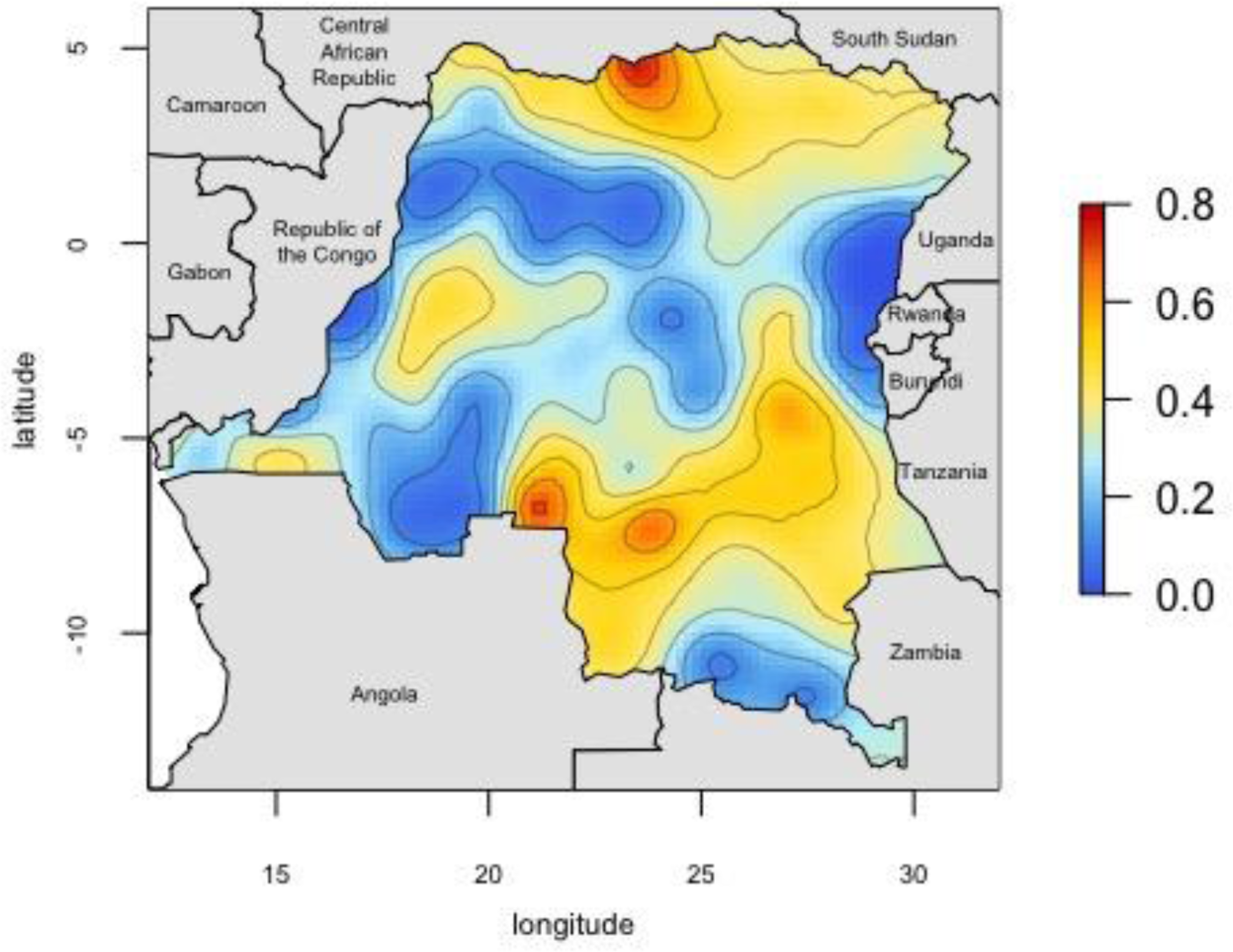
Map of predicted *P. falciparum* PCR prevalence estimates. Smoothed prevalence estimates, incorporating the sampling weights, were generated using the *PrevMap* package in R^39^. Predicted proportions range from 0-0.76.

Reported bednet use was low; though 75% of adults reported owning a LLIN, only 54% reported sleeping under a net the previous night. The average ratio of nets per household member was 0.27, indicating approximately one net per four household members. Only 17% of adults lived in houses with at least one net per two household members. Baseline characteristics of study participants are presented in **Table 1**.

### Risk factor analysis

Several covariates were associated with prevalence of malaria infection (**Table 2**). Individual protective factors included: increasing age, female sex, increasing education, and increasing wealth index. Protective household factors included living in a house made of modern housing materials and having a metal roof. At the cluster-level, increasing median wealth index was protective. Higher use of SP amongst pregnant women at the cluster level was also found to be associated with lower prevalence of malaria. Drug-resistance mutations were more common in low-prevalence clusters. Increasing average temperature was associated with increased prevalence of infection. Full risk factor modeling results are available in Table 2.

**Table 2:**
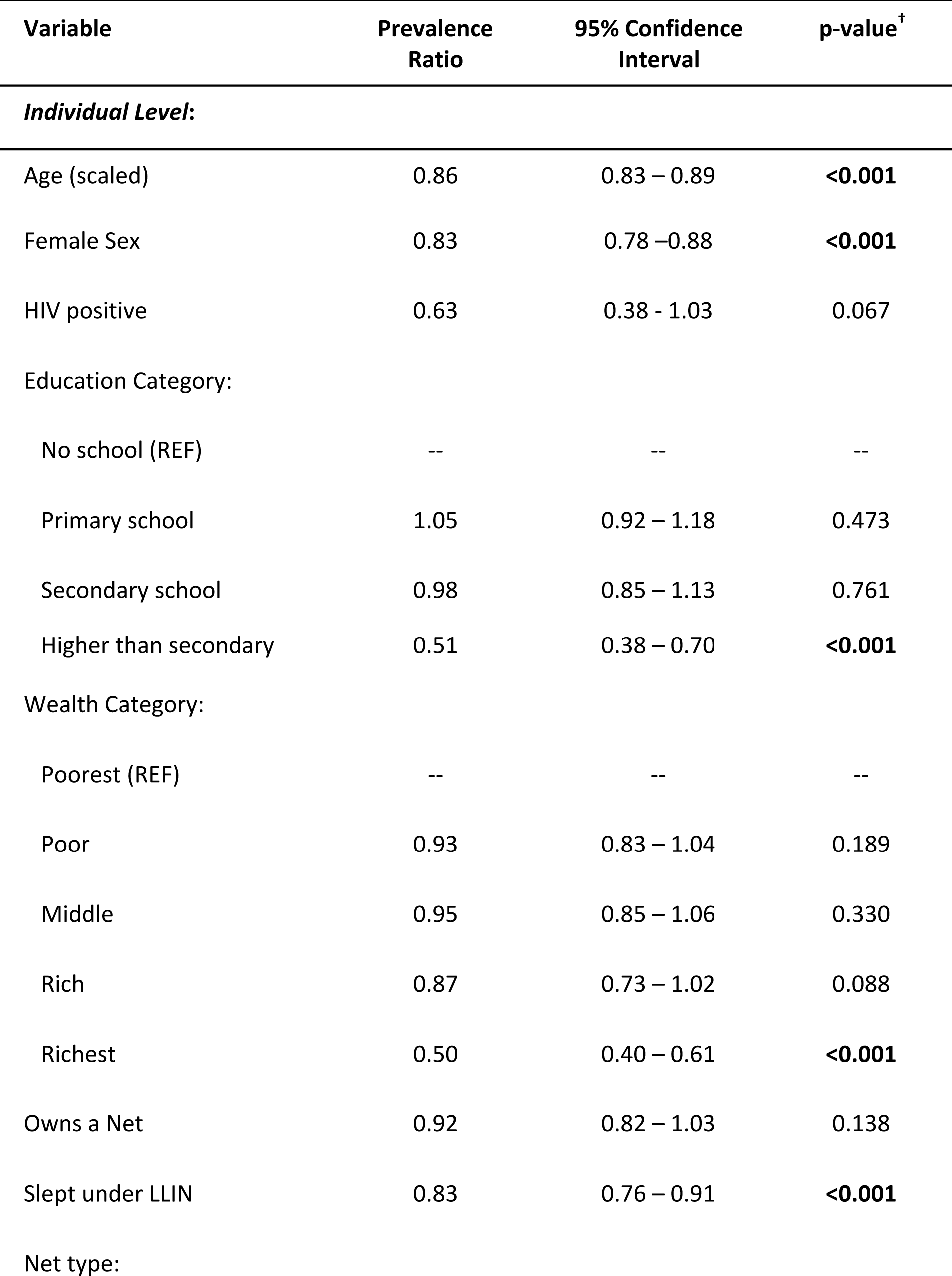
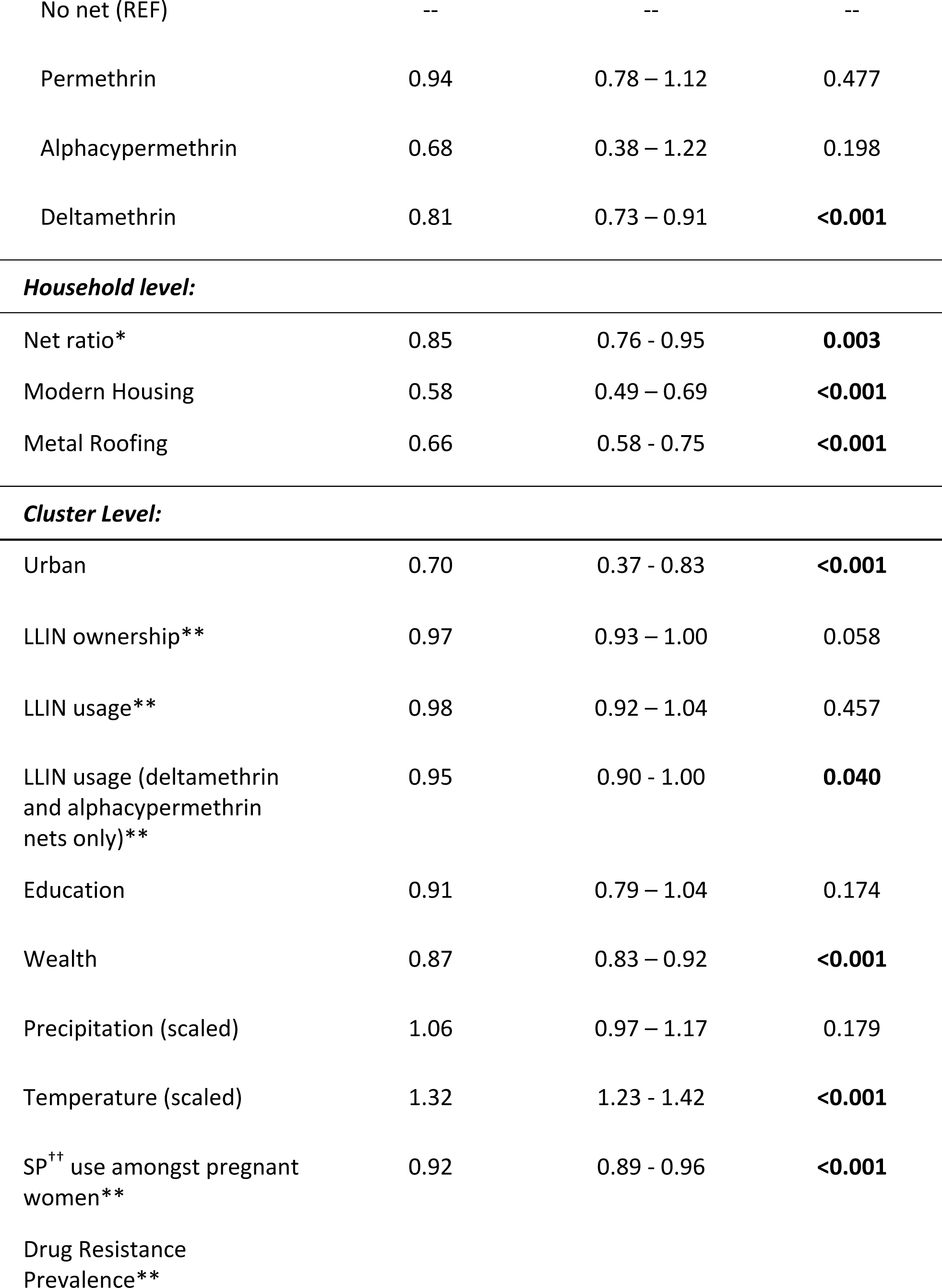
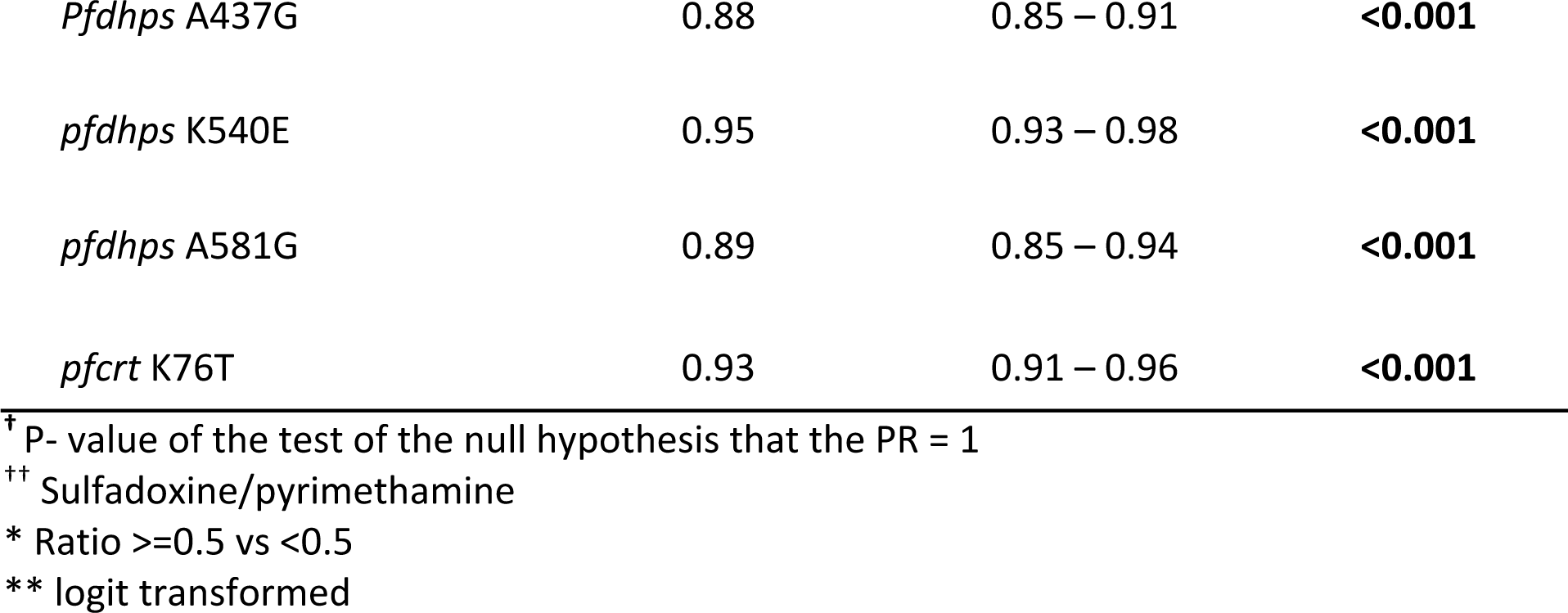
Risk factor analysis results

| Variable | Prevalence Ratio | 95% Confidence Interval | p-value<sup>†</sup> |
| --- | --- | --- | --- |
| *Individual Level:* |  |  |  |
| Age (scaled) | 0.86 | 0.83 – 0.89 | **<0.001** |
| Female Sex | 0.83 | 0.78 – 0.88 | **<0.001** |
| HIV positive | 0.63 | 0.38 - 1.03 | 0.067 |
| Education Category: |  |  |  |
| No school (REF) | -- | -- | -- |
| Primary school | 1.05 | 0.92 – 1.18 | 0.473 |
| Secondary school | 0.98 | 0.85 – 1.13 | 0.761 |
| Higher than secondary | 0.51 | 0.38 – 0.70 | **<0.001** |
| Wealth Category: |  |  |  |
| Poorest (REF) | -- | -- | -- |
| Poor | 0.93 | 0.83 – 1.04 | 0.189 |
| Middle | 0.95 | 0.85 – 1.06 | 0.330 |
| Rich | 0.87 | 0.73 – 1.02 | 0.088 |
| Richest | 0.50 | 0.40 – 0.61 | **<0.001** |
| Owns a Net | 0.92 | 0.82 – 1.03 | 0.138 |
| Slept under LLIN | 0.83 | 0.76 – 0.91 | **<0.001** |
| Net type: |  |  |  |
| No net (REF) | -- | -- | -- |
| Permethrin | 0.94 | 0.78 – 1.12 | 0.477 |
| Alphacypermethrin | 0.68 | 0.38 – 1.22 | 0.198 |
| Deltamethrin | 0.81 | 0.73 – 0.91 | **<0.001** |
| ***Household level:*** |  |  |  |
| Net ratio* | 0.85 | 0.76 - 0.95 | **0.003** |
| Modern Housing | 0.58 | 0.49 – 0.69 | **<0.001** |
| Metal Roofing | 0.66 | 0.58 - 0.75 | **<0.001** |
| ***Cluster Level:*** |  |  |  |
| Urban | 0.70 | 0.37 - 0.83 | **<0.001** |
| LLIN ownership** | 0.97 | 0.93 – 1.00 | 0.058 |
| LLIN usage** | 0.98 | 0.92 – 1.04 | 0.457 |
| LLIN usage (deltamethrin and alphacypermethrin nets only)** | 0.95 | 0.90 - 1.00 | **0.040** |
| Education | 0.91 | 0.79 – 1.04 | 0.174 |
| Wealth | 0.87 | 0.83 – 0.92 | **<0.001** |
| Precipitation (scaled) | 1.06 | 0.97 – 1.17 | 0.179 |
| Temperature (scaled) | 1.32 | 1.23 - 1.42 | **<0.001** |
| SP<sup>++</sup> use amongst pregnant women** | 0.92 | 0.89 - 0.96 | **<0.001** |
| Drug Resistance Prevalence** |  |  |  |
| *Pfdhps* A437G | 0.88 | 0.85 – 0.91 | **<0.001** |
| *pfdhps* K540E | 0.95 | 0.93 – 0.98 | **<0.001** |
| *pfdhps* A581G | 0.89 | 0.85 – 0.94 | **<0.001** |
| *pfCRT* K76T | 0.93 | 0.91 – 0.96 | **<0.001** |

<sup>†</sup> P- value of the test of the null hypothesis that the PR = 1
<sup>††</sup> Sulfadoxine/pyrimethamine
\* Ratio $\geq 0.5$ vs $< 0.5$
\*\* logit transformed

We found a modest protective association of individual LLIN use and no effect of increasing community LLIN use, in contrast to previous findings in the DRC^9^. At the household level, a net ratio greater than 0.5 (i.e. at least one net per two household individuals) was protective, compared to households with a lower ratio. When we stratified individual net use by insecticide type, we observed a protective effect of deltamethrin-treated nets and a signal for protection with alphacypermethrin nets. There was no significant protective effect of permethrin-treated nets, a finding that has been previously reported for children in the DRC^9^. Restricting LLIN use to only those who had reported using a net treated with deltamethrin or alphacypermethrin, we found a slight protective effect of increasing community level net use. The sensitivity analysis of net use showed no differences between measurement options or the coding choice for net use (**Supplemental Figure 1**). A comparison of individual LLIN net use between adults and children included in the 2013-2014 DHS indicated similar protective effects (**Supplementary Table 2**).

### Analysis of Associations by Urban/Rural status

There were several socioeconomic-related risk factors whose association varied by urban/rural status (**Figure 4**). These included individual level wealth, with increasing wealth showing a protective effect in urban areas but not rural. A similar trend was observed for increasing individual level education. Additionally, both modern housing and metal roofs were protective in urban areas but not in rural areas. Cluster level wealth demonstrated problematically high collinearity with urbanicity and thus could not be modeled. Full numeric results from this analysis are presented in **Supplemental Table 1**.

**Figure 4:**
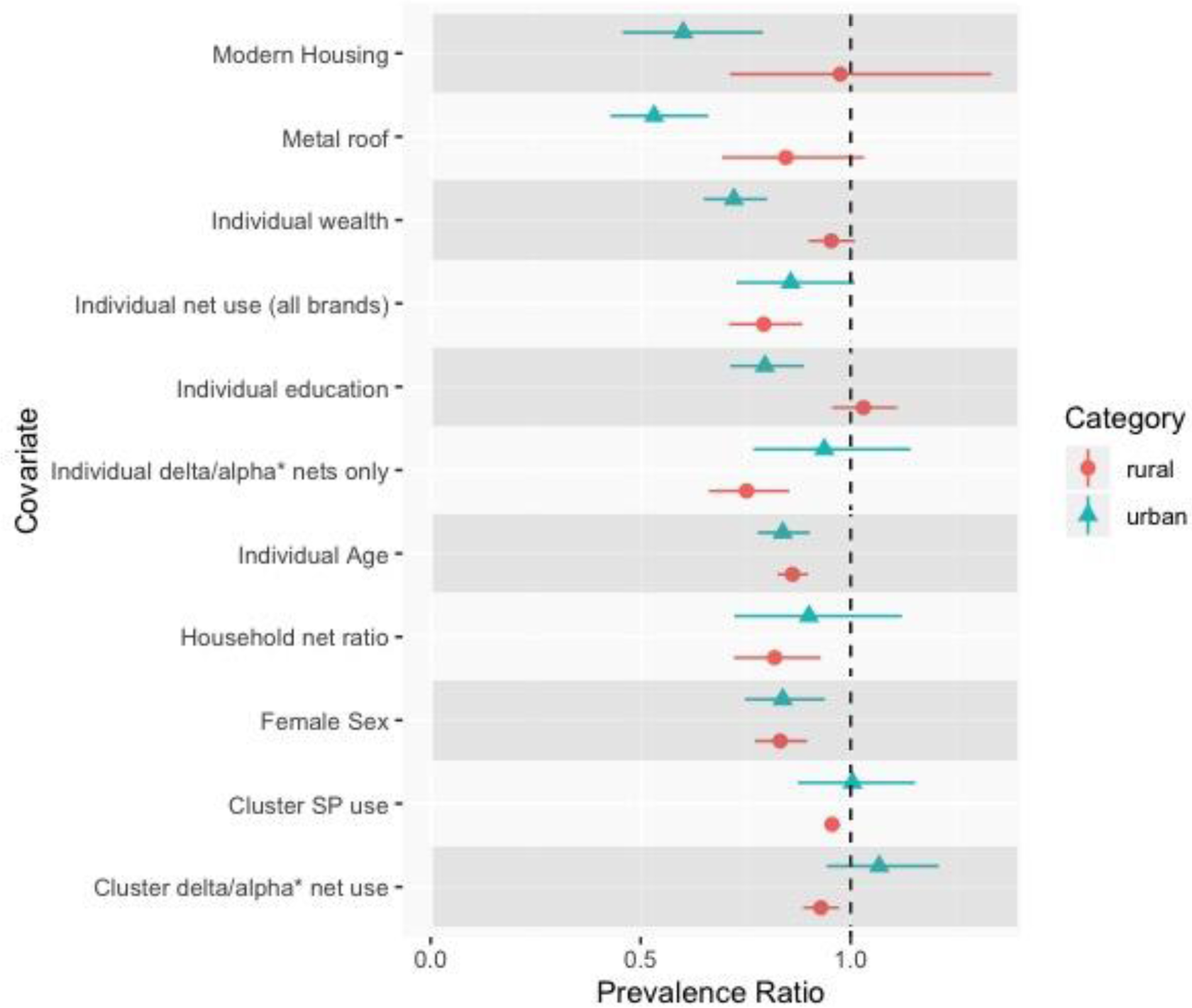
Results of the analysis comparing risk factor effects between urban versus rural areas. Prevalence ratios and confidence intervals by urban status are presented for each risk factor. Urban results are presented with blue triangles, rural results with red circles. Differences in point estimates indicate differences of the prevalence ratio by urbanicity. The associations of several factors, such as modern housing, education, and wealth, demonstrated differences between urban and rural areas. The null value (Prevalence Ratio = 1) is indicated with a vertical dashed line. *deltamethrin and alphacypermethrin net use only

Overall, urbanicity did not impact the association between LLIN use and malaria prevalence, either at the individual or cluster level. However, when we subset LLIN use to only deltamethrin and alphacypermethrin nets, the only insecticides that demonstrated a protective effect in the initial analyses, we observed that increasing individual and community use of these nets were more protective in rural areas than in urban areas. Similarly, the analysis of LLIN use by cluster-level prevalence found that individual net use was more protective in areas of higher overall malaria prevalence (**Figure 5**).

**Figure 5:**
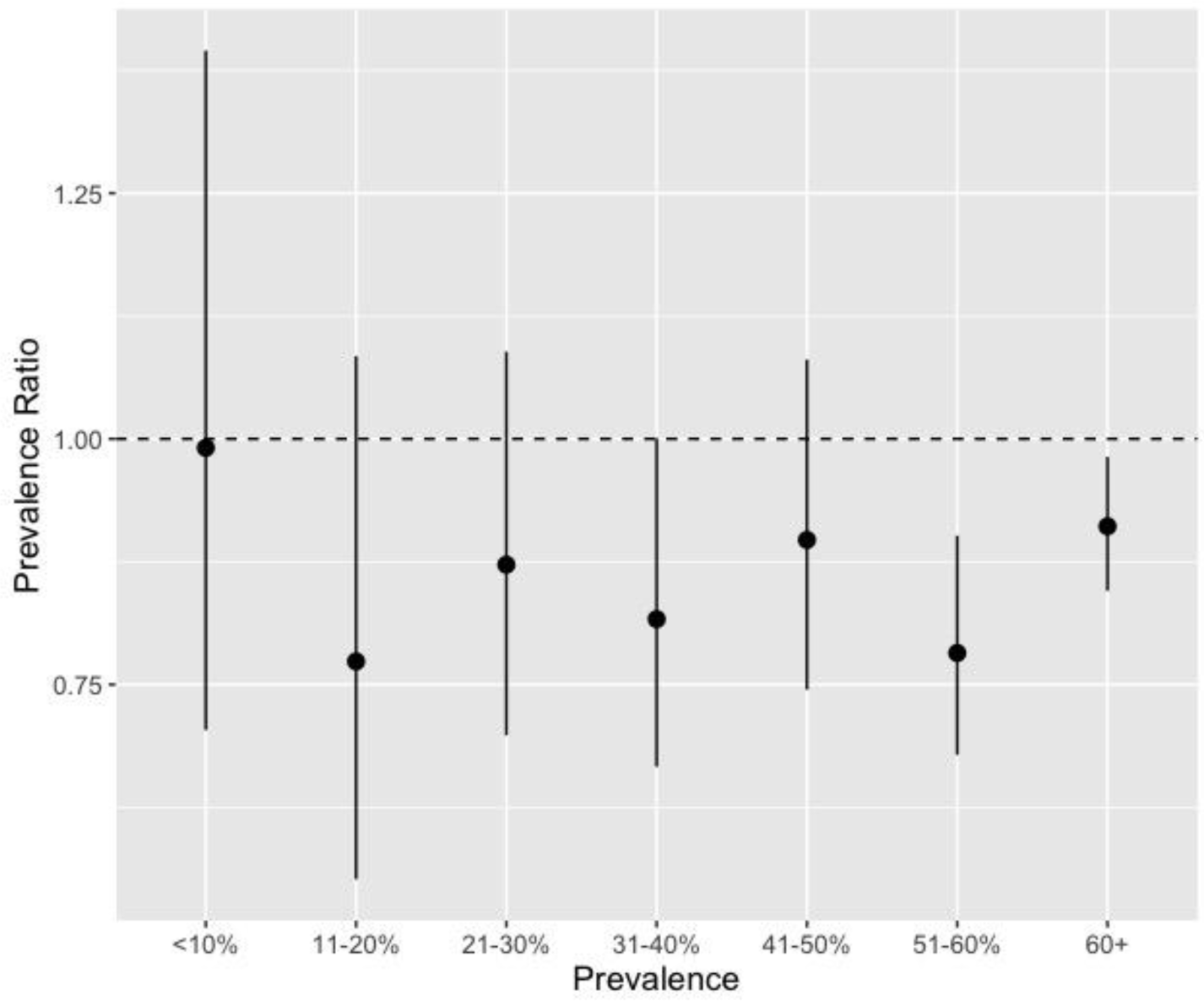
Effect of individual bednet use by cluster-level malaria prevalence. We used previously published data from children included in the 2013-2014 Demographic and Health Survey in order to determine cluster-level prevalence^28^. Individual bednet use was more protective in clusters with higher malaria prevalence. The null value (Prevalence Ratio = 1) is indicated with a horizontal dashed line.

## Discussion

In this study, utilizing data from the largest and most recent nationally-representative health survey conducted in the DRC, we found a high national prevalence of *P. falciparum* malaria (30%) amongst adults. We also found high spatial heterogeneity. Community level prevalence estimates reached as high as 76%. Malaria is often considered a pediatric disease as the majority of deaths occur amongst children^5^. However, our findings emphasize that the burden of disease amongst adults is high in the DRC and highlight the importance of adult infections in malaria epidemiology. The high level of spatial heterogeneity across the country underscores the need to study malaria on a sub-national scale. The spatial distribution of infections amongst adults matches that of infections amongst children included in the 2013-2014 DHS, indicating similar underlying spatial processes^49^.

The national prevalence estimate of 30% amongst adults is similar to that from the 2007 DHS, suggesting a need for increased investment in malaria control in the DRC and further understanding of drivers of transmission^1,13^. We observed that the prevalence is unchanged from 2007 despite increases in LLIN coverage and the number of individuals being tested and treated for infection^50^. Thus, understanding malaria risk factors in all age groups remains critical for the design and implementation of effective interventions. Several risk factors identified in an analysis of the 2007 DHS data were also associated with increased infection prevalence in this study, highlighting their continued importance in the epidemiology of malaria. These include younger age, male sex, and lower community level wealth^13^. The factors identified in this study, many of which have also been identified in other countries^51–53^, are particularly important for identifying infected individuals in the DRC, as many adult infections are asymptomatic and often go undetected^8^. Thus, targeting malaria interventions towards younger adults, men, and poorer communities could maximize their impact.

In this study, we found that individuals living in households with a bednet ratio greater than 0.5, ie: at least one net for every two household members, had lower prevalence of malaria. This ratio is recommended by the WHO in order to ensure a sufficient number of nets for household members^54^. Though previous studies have assessed the role of increasing community level net use, the effect of within household “net coverage” has not been studied, to our knowledge. Our findings demonstrate the importance the number of nets within a household and support the WHO recommendation. However, less than 20% of survey respondents lived in households with a net ratio of 0.5 or higher. Thus, the DRC is far from achieving the WHO recommendation. It is critical that future bednet distribution campaigns ensure that enough nets are provided to each household.

In this analysis we did not observe the overall protective effect of community bednet use that has been previously demonstrated amongst children in the DRC, though there was a protective effect of increasing community use of deltamethrin and alphacypermethrin treated nets^9^. As mentioned, we did see a protective effect of increasing within household net coverage, which may be a more important factor for reducing infection than community net use. Our findings of greater protection from nets in higher prevalence areas and in rural areas are supported by a recent meta-analysis that found increasing community malaria prevalence associated with a greater protective effect of LLIN use (OR = 0.80)^55^. Thus, bednet distribution campaigns may be more successful in rural or generally higher prevalence areas than in cities.

The findings from the genetic analysis of mutations associated with SP and chloroquine resistance indicate that areas of higher prevalence of drug resistance had lower prevalence of malaria infection. While this is a marginal association and does not reflect a causal relationship, this could be due to the amount of anti-malarial drug use, leading to lower overall prevalence of malaria but increased resistance through increased selective pressure. Future studies should aim to investigate the causal effect of increasing community level drug resistance on malaria risk; however, this was not the primary aim of this study. Additionally, cluster-level SNP prevalences were modeled estimates generated using data from children and thus may not be representative of overall community level prevalence.

Modern housing and metal roofing were both associated with lower prevalence of malaria, though the association was more pronounced in urban areas than in rural areas. The effect of metal roofing agrees with findings from a recent study conducted in the Gambia, which found 38% lower mosquito survival and lower malaria prevalence amongst villages with higher proportions of metal roofs^56^. The authors of the study propose that this is due to the higher temperatures of metal roofs during the day, leading to lower mosquito survival. The present findings suggest that housing improvements may help reduce malaria risk, either directly as proposed by the Gambian study, or indirectly through overall improvements in living conditions and socioeconomic status.

This study also highlights differences in the epidemiology of malaria between urban and rural areas, confirming the need to tailor interventions to different populations in cities versus rural areas. Increasing wealth and education are both highly protective in urban areas but not in rural areas. This may be a result of increased access to prevention and treatment methods in urban areas, as has been observed in other countries. A study from Equatorial Guinea found individuals in rural areas waited longer to seek treatment for malaria infections compared to those in urban areas, and were more likely to be treated at home rather than in a health facility^57^. These results also agree with findings from previous studies that poor individuals living in urban areas have similar health risks to the general rural population^58^. Thus, in urban areas, the malaria positive population is poorer and less educated than those infected in rural areas. Malaria control programs should take these differences in high-risk populations into account, targeting poorer or less educated individuals in cities and ensuring that interventions are accessible for these populations. Conversely, interventions in rural areas need to be more widespread and accessible to a more diverse population.

This study has several strengths. First, it uses nationally-representative, population-based data from over 16,000 individuals, the largest health survey conducted in the DRC. This allows us to make inferences regarding the country as a whole. Other recent malaria risk factor studies have fewer individuals or are conducted in a smaller geographic area^11,12^. Second, we determined malaria infection using high-throughput real-time PCR, the most sensitive method for diagnosing infection^28^. Third, it leverages country-wide drug resistance genotyping data to inform community-level epidemiological analysis. Lastly, this study incorporates multiple types of data including: survey, molecular, deep sequencing drug resistance, and geospatial data collected at the individual, household, community, and environmental level. This allowed us to demonstrate the different scales of malaria risk factors; control programs should aim to intervene at each of these levels.

The findings from this study are subject to limitations. Several of the covariates included in the study were obtained from self-reports, including LLIN usage. While self-reported data is subject to recall bias, the DHS questionnaire asks multiple questions regarding net use that allowed us to assess bias. The sensitivity analysis comparing these questions indicated no difference in modeling results between the questions, providing confidence that bias from self-reports is minimal. Secondly, this analysis evaluated marginal associations and thus the findings cannot be interpreted as causal effects. However, the associations are useful for identifying and targeting interventions to higher risk individuals and groups such as younger, less educated men. Additionally, as the DHS is a cross-sectional survey, we did not assess the effects of seasonality. Finally, we could not directly assess the epidemiology of symptomatic malaria in the DRC as DHS surveys do not sample health facilities.

## Conclusion

This study evaluated risk factors for *P. falciparum* infection amongst adults in the DRC using data from the nationally representative, population based DHS survey. Overall prevalence of infection was high, 30%, and grossly unchanged from the prior 2007 survey. We observed high spatial heterogeneity across the country and identified individual, household, and community-level risk factors for malaria such as male sex, modern housing, and increasing within household net coverage. These findings support the need for sustained investment in malaria control in the DRC and can be used to develop targeted interventions with maximal impact.

## Data Availability

Data used in this study from the Demographic Health Surveys Program are available upon request at (https://dhsprogram.com/what-we-do/survey/survey-display-421.cfm)59. Raw sequencing data is publicly available through the NCBI SRA (Ascension number: PRJNA545347). Molecular data included in this study are available from the corresponding author upon reasonable request.

https://dhsprogram.com/what-we-do/survey/survey-display-421.cfm

## List of Abbreviations

DRC: Democratic Republic of the Congo
DHS: Demographic and Health Survey
PCR: polymerase chain reaction
LLIN: long-lasting insecticide treated net
SNP: single nucleotide polymorphism
MIP: molecular inversion probe
PCA: principal components analysis
SP: Sulfadoxine/pyrimethamine

## DISCLOSURES

### Ethics Approval and Consent to Participate

This study was approved by the Internal Review Board at The University of North Carolina, Chapel Hill and at the Kinshasa School of Public Health. Informed consent was obtained from each individual age 18 or older, or from a parent or legal guardian for children and adolescents under age 18.

## Consent for Publication

Not Applicable

## Availability of Data and Materials

Data used in this study from the Demographic Health Surveys Program are available upon request at (https://dhsprogram.com/what-we-do/survey/survey-display-421.cfm)^59^. Raw sequencing data is publicly available through the NCBI SRA (Ascension number: PRJNA545347). Molecular data included in this study are available from the corresponding author upon reasonable request.

## Author Contributions

MDF conducted the analyses and drafted the manuscript. NFB provided assistance with data management and the risk factor analyses. JBP and KLT conducted the PCR assays. JM supervised blood spot processing. MK and AT oversaw sample management. OA and JAB generated and processed the sequencing data. JE, RV, ME, and EG provided guidance with the study design and provided analytical assistance. JJJ and SRM conceived of the study and oversaw study set up. All authors edited the manuscript and approved the final draft.

## Acknowledgements

The authors would like to thank all the study participants and all study team members. The authors would also like to thank Stephanie Doctor for assistance with sample processing.

## Supplementary Text

### Modern Housing variable construction

We constructed a composite “modern housing” variable using data from the roof, wall, and floor material variables reported in the DHS. The definition of modern housing was based off those used by Tusting et al^1^. Modern roofing materials included: metal, zinc/cement, tiles/slate, or cement. Modern wall material included: cement, stone, bricks, or covered adobe. Modern floor materials included: vinyl, asphalt, ceramic tiles, cement, or carpet. Only houses with a modern roof, walls, and floor materials were considered “modern housing”.

### PrevMap analysis

We estimated cluster-level drug resistance allele frequencies using the *PrevMap* package in R^2^. We fit a model in order to generate cluster-level SNP prevalence estimates at all sampled DHS clusters from the 1,065 children with available data. Each resistance mutation was analyzed individually. We first determined raw cluster-level SNP frequencies and then transformed the proportions using a logit transformation. We fit linear a geospatial model of the following form:

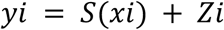

In this model,*yi* is equal to the transformed allele frequency for each cluster and *S* (*xi*) represents an isotropic Gaussian Process with a variance of ***σ***^2^ and a Matern correlation function^2^. *Zi* represents a Gaussian error term^2^. Model parameters were estimated using maximum likelihood and models were run using 10,000 simulations to generate spatially smooth frequency estimates. After fitting the model, we extracted the frequency estimate for each DHS cluster included in the analysis. To minimize bias, we averaged the estimated values for all geopoints within 15km square of the DHS cluster geopoint.

We used the same model framework to generate spatially smoothed *P. falciparum* prevalence estimates (**Figure 3**). Using this model, we also determined the estimate error for all points (**Supplementary Figure 1**).

### Bednet use sensitivity analysis

We evaluated four different methods for determining bednet usage based on the questions asked in the DHS questionnaire. The first (M1) asked if the individual slept under an “ever treated” net the previous night. The second (M2) asked if the individual slept under a “long lasting insecticide treated net”. For the third method (M3), we constructed a “new net” variable based on whether the individual had obtained the net, or re-treated it with insecticide, within the previous 3 years^1^. Lastly (M4), the DHS asks, “the type of mosquito bednet person slept under”, with options of no net, an untreated net, or a treated net. We compared the estimated prevalence ratio of those who used nets versus those who did not based on each method’s definition of net use. The results (**Figure S2**) demonstrate no substantial differences in the effect of net use between the four methods. The variable used in M1 was used for the primary analysis.

## Supplementary Tables/Figures

**Figure S1:**
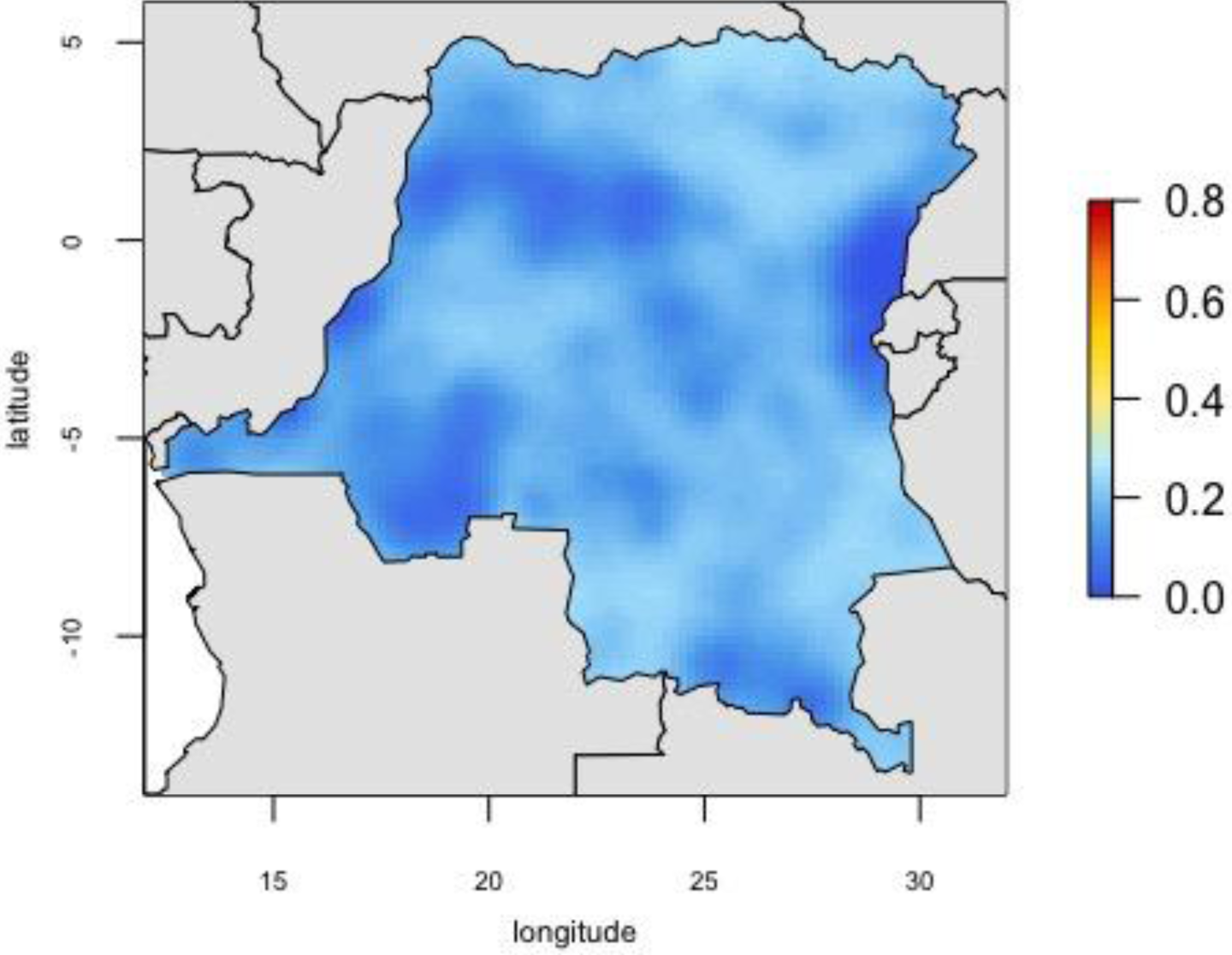
Standard error map for smoothed *P. falciparum* PCR prevalence estimates generated using *PrevMap*^2^.

**Figure S2:**
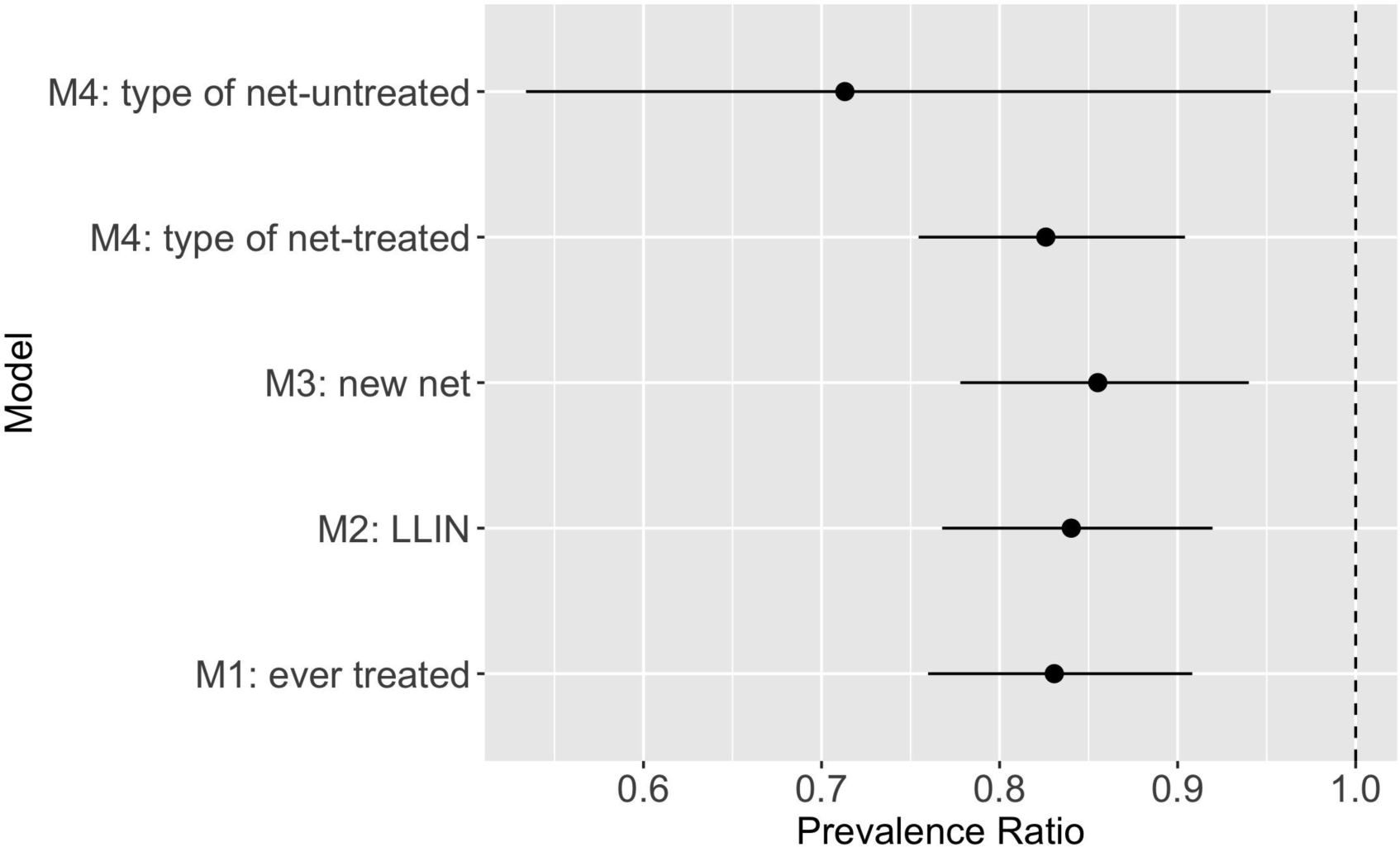
Results of sensitivity analysis evaluating different methods for measuring and coding bednet usage. Each model compared the prevalence of PCR detectable *P. falciparum* amongst individuals who reported using a net versus those who did not.

**Table S1:** Province level *P*.*falciparum* prevalence estimates measured by PCR.

| **Province name** | **Prevalence %**<br>**(SE)** |
| --- | --- |
| Kinshasa | 18.6 (2.3) |
| Kwango | 21.4 (3.7) |
| Kwilu | 25.8 (5.0) |
| Mai-Ndombe | 43.9 (4.7) |
| Kongo Central | 40.5 (3.8) |
| Equateur | 22.4 (2.7) |
| Mongala | 26.3 (2.9) |
| Nord-Ubangi | 48.9 (5.0) |
| Sud-Ubangi | 31.3 (5.3) |
| Tshuapa | 33.7 (4.4) |
| Kasai | 44.5 (7.3) |
| Kasai-Central | 43.5 (4.0) |
| Kasai-Oriental | 35.9 (4.3) |
| Lomami | 52.5 (3.1) |
| Sankuru | 27.2 (3.3) |
| Haut-Katanga | 22.5 (3.4) |
| Haut-Lomami | 41.0 (5.2) |
| Lualaba | 38.6 (9.6) |
| Tanganyika | 54.3 (5.1) |
| Maniema | 45.5 (5.7) |
| Nord-Kivu | 6.7 (1.6) |
| Bas-Uele | 58.3 (6.4) |
| Haut-Uele | 42.7 (3.6) |
| Ituri | 33.9 (4.1) |
| Tshopo | 29.6 (4.9) |
| Sud-Kivu | 16.4 (3.2) |

**Table S2:**
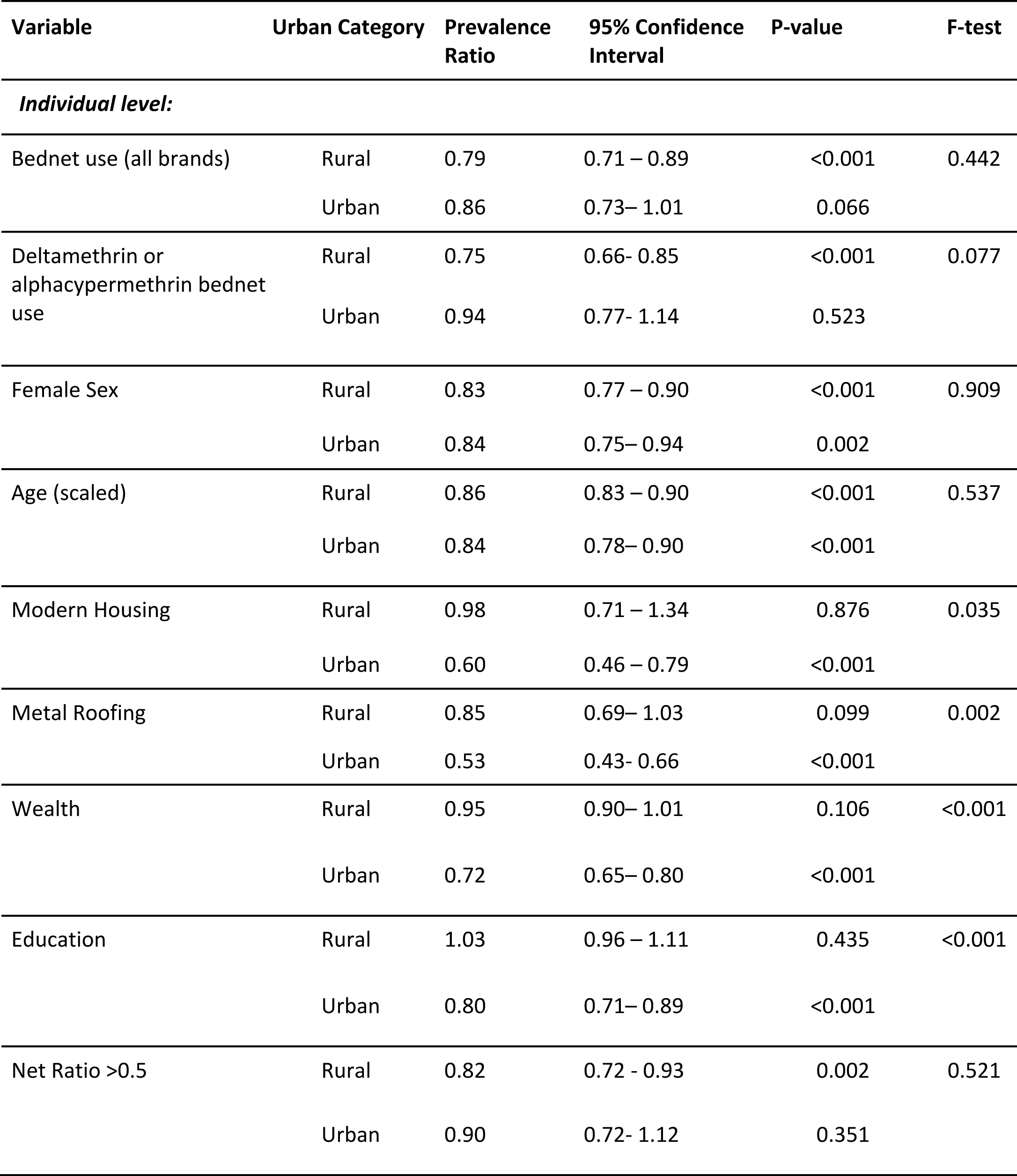
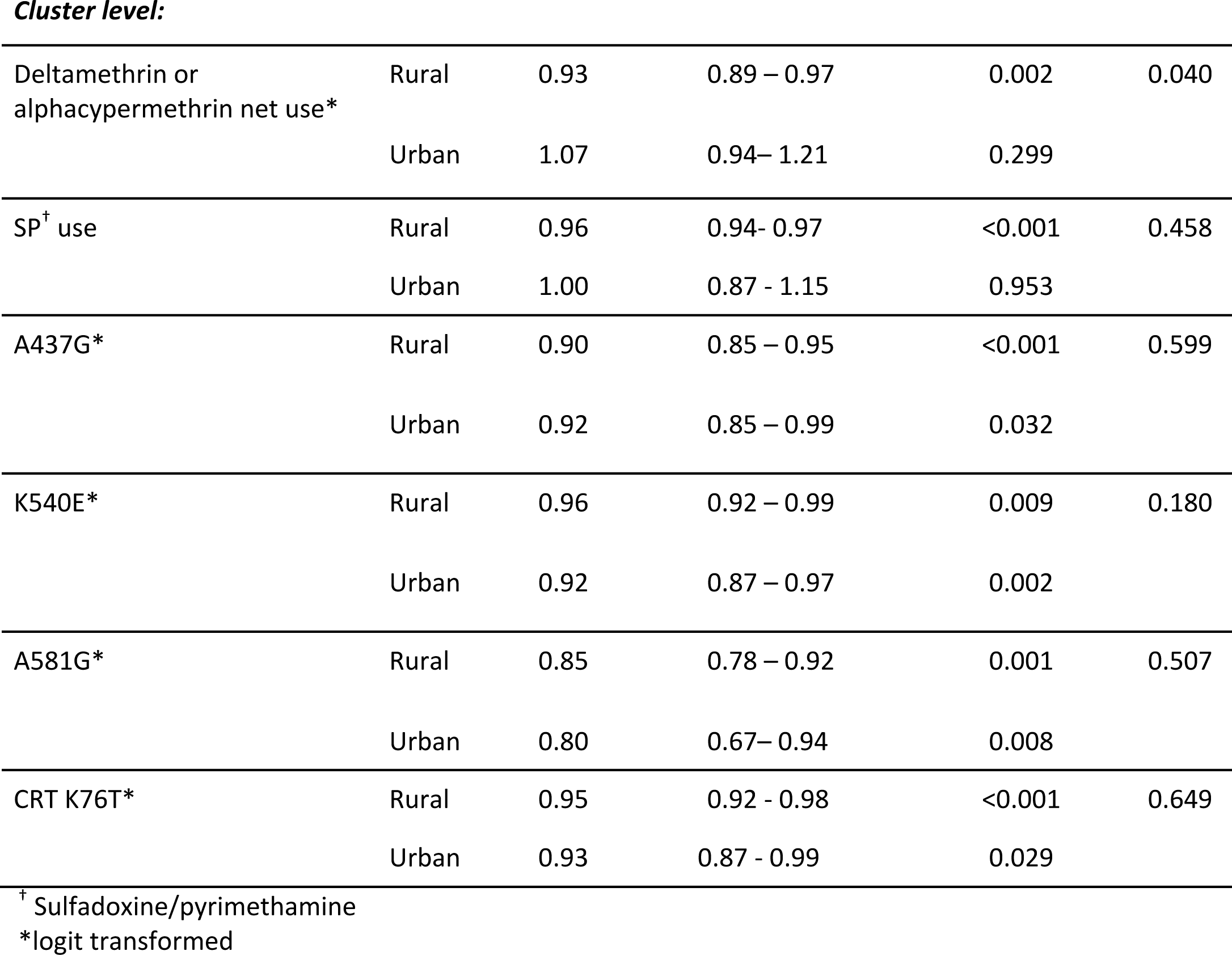
Associations between identified risk factors and *P. falciparum* prevalence, stratified by urbanicity

| Variable | Urban Category | Prevalence Ratio | 95% Confidence Interval | P-value | F-test |
| --- | --- | --- | --- | --- | --- |
| *Individual level:* |  |  |  |  |  |
| Bednet use (all brands) | Rural | 0.79 | 0.71 – 0.89 | <0.001 | 0.442 |
|  | Urban | 0.86 | 0.73– 1.01 | 0.066 |  |
| Deltamethrin or alphacypermethrin bednet use | Rural | 0.75 | 0.66- 0.85 | <0.001 | 0.077 |
|  | Urban | 0.94 | 0.77- 1.14 | 0.523 |  |
| Female Sex | Rural | 0.83 | 0.77 – 0.90 | <0.001 | 0.909 |
|  | Urban | 0.84 | 0.75– 0.94 | 0.002 |  |
| Age (scaled) | Rural | 0.86 | 0.83 – 0.90 | <0.001 | 0.537 |
|  | Urban | 0.84 | 0.78– 0.90 | <0.001 |  |
| Modern Housing | Rural | 0.98 | 0.71 – 1.34 | 0.876 | 0.035 |
|  | Urban | 0.60 | 0.46 – 0.79 | <0.001 |  |
| Metal Roofing | Rural | 0.85 | 0.69– 1.03 | 0.099 | 0.002 |
|  | Urban | 0.53 | 0.43- 0.66 | <0.001 |  |
| Wealth | Rural | 0.95 | 0.90– 1.01 | 0.106 | <0.001 |
|  | Urban | 0.72 | 0.65– 0.80 | <0.001 |  |
| Education | Rural | 1.03 | 0.96 – 1.11 | 0.435 | <0.001 |
|  | Urban | 0.80 | 0.71– 0.89 | <0.001 |  |
| Net Ratio >0.5 | Rural | 0.82 | 0.72 - 0.93 | 0.002 | 0.521 |
|  | Urban | 0.90 | 0.72- 1.12 | 0.351 |  |
| **Cluster level:** |  |  |  |  |  |
| Deltamethrin or<br>alphacypermethrin net use* | Rural | 0.93 | 0.89 – 0.97 | 0.002 | 0.040 |
|  | Urban | 1.07 | 0.94– 1.21 | 0.299 |  |
| SP<sup>†</sup> use | Rural | 0.96 | 0.94- 0.97 | <0.001 | 0.458 |
|  | Urban | 1.00 | 0.87 - 1.15 | 0.953 |  |
| A437G* | Rural | 0.90 | 0.85 – 0.95 | <0.001 | 0.599 |
|  | Urban | 0.92 | 0.85 – 0.99 | 0.032 |  |
| K540E* | Rural | 0.96 | 0.92 – 0.99 | 0.009 | 0.180 |
|  | Urban | 0.92 | 0.87 – 0.97 | 0.002 |  |
| A581G* | Rural | 0.85 | 0.78 – 0.92 | 0.001 | 0.507 |
|  | Urban | 0.80 | 0.67– 0.94 | 0.008 |  |
| CRT K76T* | Rural | 0.95 | 0.92 - 0.98 | <0.001 | 0.649 |
|  | Urban | 0.93 | 0.87 - 0.99 | 0.029 |  |

<sup>†</sup> Sulfadoxine/pyrimethamine
\*logit transformed

**Table S3:** Comparison of the association between individual LLIN use vs no LLIN^†^ use and malaria prevalence between adults and children in the 2013-2014 Demographic and Health Survey. Data from children has been previously published^3,4^.

| Population | Prevalence Ratio | 95% Confidence Interval |
| --- | --- | --- |
| Children | 0.82 | 0.72 – 0.91 |
| Adults | 0.83 | 0.76 – 0.91 |

<sup>†</sup> Long-lasting insecticide treated net

